# TAMING COVID-19 EPIDEMIC IN SÃO PAULO WITH A LOGISTIC MODEL AND NON-PHARMACEUTICAL MEASURES

**DOI:** 10.1101/2020.05.06.20093666

**Authors:** Marcelo Marchesin

**Author notes:** **AMS** : 92D30, 93C15, 34H05.

## Abstract

In this paper I use the simple logistic mathematical model to represent the development of COVID-19 epidemic in São Paulo city under quarantine regime and I estimate the total amount of time it is necessary to decrease the number of seriously ill patients in order to reduce the demand for hospital beds of Intensive Care Units (ICU) to tolerable levels. Clearly the same reasoning and mathematical methods used in here can be used for any other city in similar conditions of social isolation.

## 1. Introduction

On December 2019 in Wuhan City (China), several cases of pneumonia of unknown etiology were detected. The Chinese Country Office of the World Health Organization was informed and a novel coronavirus (officially named SARS-Cov-2) was identified on January 7*^th^*, as the cause of such infection. The disease was officially named COVID-19. An international alert was issued because an eminent potential for worldwide spread had been recognized.

According to the website Worldometers [10] which features the first figures on China (Wuhan), on January 22*^nd^* there were already 571 confirmed cases and 2 days later there were already 1, 287 with 41 deaths. COVID-19 has been shown to be easily spreading and very lethal. China’s effort to mitigate the harm were quickly taken yet as many as more than 75, 000 infected cases were reached in Wuhan before the end of January (see [3]). Due to the highly interconnected world we presently live in, the disease quickly spread outside China reaching practically all countries around the world with several different degrees of seriousness. On March 11*^th^* due to the worrying situation the World Health Organization (WHO) declared it a Pandemic.

The scientific world community understood it was time for an international effort to provide scientific reliable information to help the world’s leaders in the choice of public policies to face the pandemic. On March 16*^th^* a group of about 30 researchers from the London Imperial College published a blunt paper [2] considering the various options of several different public policies measures that should be taken. They have analyzed several possibilities varying from total no intervention to complete lock-down of the whole population and they have estimated the number of lives loss in each case. Lately, because of the seriousness of the situation, there has been many other papers researching the same topic: Li Q. et. al. [3],WuP. et. al.[14],Wu J. et. al.[13], Rocha Filho T.M. et al.[5] just to mention a few. The last one does a nice specific study of the situation in São Paulo city.

Although the motivation for this study is the present situation of the epidemic in Brazil, we understand that the results presented here are general enough to work as a guideline to any other country and city facing the same uncertainties. Given the continental size of Brazil and the ongoing local policy of quarantine it seems reasonable to face the problem locally. At the present moment most of the Brazilian cities, specially the large ones, are fairly well isolated from the others so that a local approach is justified. I particularly study the case of São Paulo city because it was the Brazilian city where the first case of COVID-19 was diagnosed, it has been under quarantine practically since the beginning of the onset, it presents relatively reliable data on the epidemic and it is the largest and most important city in Brazil.

This paper consists of using the logistic mathematical model to the epidemic situation in São Paulo city. I study the current situation of the total number of Intensive Care Unit (ICU) beds of the public health system in São Paulo and then I use the results that the quarantine in Wuhan has produced in the decreasing number of the total number of cases of infected individuals, to infer similar results to São Paulo considering several levels of strictness of its quarantine model. I conclude estimating the amount of time necessary to decrease the number of simultaneously infected individuals in order to avoid overwhelming the ICU public health system of the city of São Paulo.

This paper is organized as follows: In Section 2, I present the mathematical model I shall use: the logistic model. In order to justify the choice of such a model, in Section 3, I fit the data from China to it to show that is has an awesome level of accuracy. In Section 4, I use the data of China (Wuhan) to estimate quantitatively how the strictness of the quarantine regime influences on the variation of the parameters *r* and *T*. In Section 5, I fit the logistic model to the data of São Paulo and finally in Section 6, I estimate when the demand for ICU beds reaches 80% of the total number of such available beds of the public health system and how long it takes to reduce these numbers depending on the level of accession of the quarantine regime in the city. In Section 7, I summarize the previous results stressing how an specific level of accession to the quarantine regime can influence in a relative relief on the demand for ICU beds of the public health system of the city of São Paulo. The theorem used in this study is presented in Section 8.

## 2. The Logistic Model

The logistic model (see [4] for more details) has been classically used modeling population growth in a scenario in which the rate of growth is exponentially quick in the beginning, being proportional to the number of the whole population, but it losses strength as it starts getting close to an upper threshold, for instance the natural storage capacity of the environment. Here this idea seems to be very logical, for the spread of the total number of infected individuals of COVID-19 is exponentially quick, as we see it in the early days, but it is reasonable to believe it will slow down as the number of exposed susceptible individual (not yet infected), decreases. It is indeed a very simple model which can be mathematically written by an unique ordinary differential equation with an initial condition:

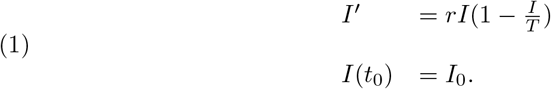

in which *t_0_* is the time chosen to represent the beginning of the outbreak, *I*_0_ in the number of infected individuals at such an initial time, *I* = *I*(*t*) is the total number of infected individuals up to time *t*, *r* is a positive constant of proportion and *T* is the total number of susceptible exposed individuals. It is said that the rate of change in the number of infected individuals is proportion to the number of infected individuals as well as to the number of remaining non-infected individuals. The solution of such equation is broadly known to be:

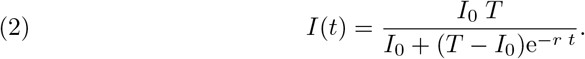

Clearly such a solution is a function of *I*_0_ as well as of *T* and *r*. We do not have any control on the number of cases at time *t* = 0 but we do have relatively great amount of control on *r* and *T* as *I* shall point out in section 6. Clearly we have that

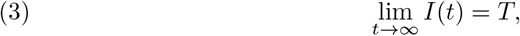

so that *T* will be the total final number of infected individuals forecasted by the model. Typical graphics of such solution functions are given in figure (1).

**Figure 1.**
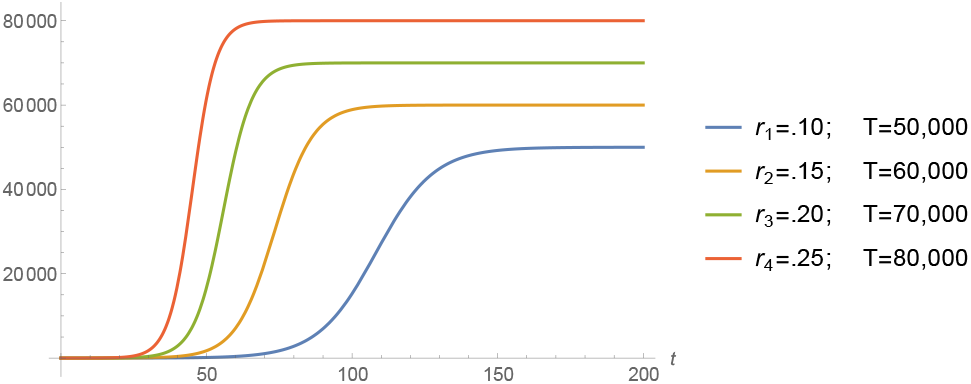
Solutions for the logistic model.

Many other papers have been written on this topic using some more sophisticated mathematical tools such as the several SIERs epidemiological models (see Castilho et. al. [1], for instance) but I have preferred to use the logistic model exactly because of its simplicity. Furthermore I shall be studying the development of the epidemic in a relatively short period of time and very locally so that such approach is easily seen to be efficient.

## 3. Fitting the Wuhan case to the Model

The most valuable argument in favor of the use of the logistic model is the extremely nice way it shows to fit the iconic “closed case” of the COVID-19 epidemic in Wuhan as we see now.

Throughout this study we shall consider the data for China, updated until May 8*^th^*. It is taken from the website Worldometer [10]. We consider the Chinese quarantine policy and we suppose the strict Chinese laws were so literally followed that the total amount of infected individual was the part of the population susceptible due to working conditions or that deliberately refused to quarantine. By April 13*^th^*, Continental China had 83, 607 confirmed cases meanwhile the Hubei province had 67, 803 confirmed cases (see [8]), mostly in its capital, Wuhan, meaning that it represents approximately 80% of all the Chinese cases.

Wuhan has a population *T*_0_ = 11,08 million people living in a region of 8.494 *Km*^2^ which features a demographic density of 1, 305 inhabitants per *Km*^2^ (see [11]). Therefore 67, 803 individuals represent less than 0.5% of its total population. Thus we can consider that the quarantine regime in Wuhan has isolated 99.50% of its whole population. Furthermore, the demographic density of the susceptible exposed individuals during the epidemic was approximately 8 individuals per *Km*^2^. That may have been the reason for the rapid success of the Chinese’s public policies.

We consider January 22*^nd^* as the date of the beginning of the outbreak in Wuhan, with initial value of 571 infected individuals. Furthermore, on April 8*^th^* Wuhan lifted the quarantine restrictions which had been imposed to its population since January 23*^rd^*, so Wuhan can be considered as the first city in the world “free of COVID-19”.

We use software Mathematica to estimate the best values for the parameters *I*_0_, *T* and *r* of the logistic model to fit China’s data. The “FindFit” tool of Mathematica, which does the fitting, is based on the least-square mathematical method to fitting date to a curve. For China it features:

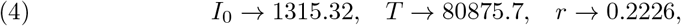

so that we get the following function for the total number of infected individuals up to time *t*, for China:

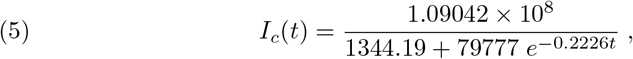

based on our previous comment we consider that the function of the total number of infected individuals for Wuhan is 80% of the one for the whole China, so we get:

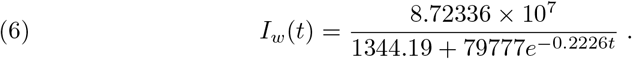

Also *r* = 0.2226 indicates the rate of infection in Wuhan. Several peculiarities for Wuhan and São Paulo (climate conditions, demographic density, advance preparation for the outbreak) would justify this rate of infection to be different in both cities. Also there is clearly a time delay between both onsets (January 22*^rd^* for Wuhan and February 25*^th^* for São Paulo), besides the difference for the initial data: 571 in Wuhan and 1 in São Paulo.

We now plot the function obtained with Mathematica together with the official data (figure 2) to see how incredibly good is the fitting for China:

**Figure 2.**
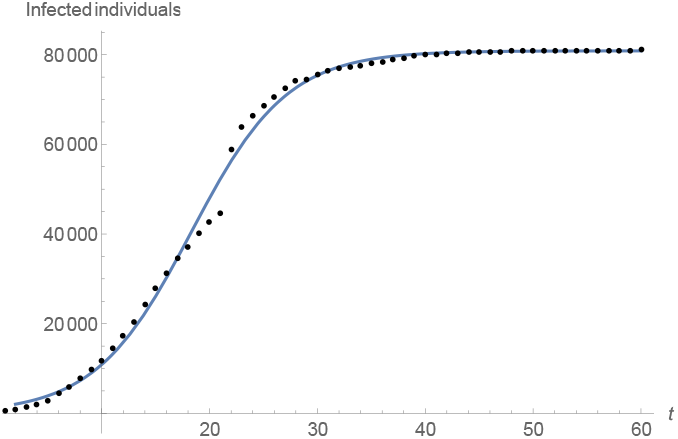
The function of the total number of infected individuals up to time *t* in China, fitting the data updated to April 21*^st^*.

**Remark:** We notice that the total number of infected individuals increased drastically on day 22 (February 12*^th^*), in China, after a change in the official methodology for diagnosing and counting cases, thousands of new cases were added to the total figures (see figure 3). We believe this will cause no problem in our reasoning.

## 4. The effects of non-pharmaceutical measures in Wuhan from an ongoing perspective

The quarantine in Wuhan was declared on January 23*^rd^*. The Chinese government closed down schools, churches and prohibited social clusters of any kind. Within about 15 days, around February 5*^th^*, the results started to be felt in the number of new cases of infected individuals (see figure (3)).

**Figure 3.**
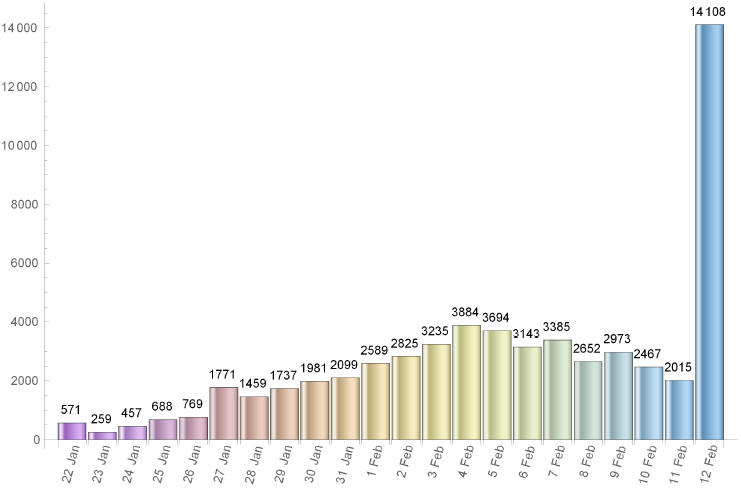
Number of new cases in China between Jan 22*^nd^* and Feb 12*^th^*.

We use the available data (from Worldometer website [10]) to estimate the impact of the quarantine measures on the estimated values of the parameters *r* and *T* obtained using Mathematical fitting tool, in several dates measured from the day 1, i.e. from January 22*^nd^*, until some days after February 5*^th^*. We present such values of *r* and *T* in the table 1

**Table 1.** Values of *r* and *T* obtained from the fitting process in several dates around the beginning of the onset.

|  | Jan 22<sup>nd</sup> | Jan 23<sup>rd</sup> | Jan 24<sup>st</sup> | Jan 25<sup>th</sup> | Jan 26<sup>th</sup> | Jan 27<sup>th</sup> | Jan 28<sup>th</sup> |
| --- | --- | --- | --- | --- | --- | --- | --- |
| $r$ | 0.4574 | 0.415475 | 0.380517 | 0.349581 | 0.319166 | 0.303118 | 0.299476 |
| $T$ | 17622 | 21548 | 26081 | 31631 | 39573 | 45218 | 46542 |
|  | Jan 29<sup>th</sup> | Jan 30<sup>th</sup> | Jan 31<sup>st</sup> | Fev 1<sup>st</sup> | Fev 2<sup>nd</sup> | Fev 3<sup>rd</sup> | Fev 4<sup>th</sup> |
| $r$ | 0.29459 | 0.29355 | 0.28922 | 0.28530 | .2825 | 0.2256 | 0.19371 |
| $T$ | 48156 | 48463 | 49596 | 50540 | 51161 | 69766 | 94572 |
|  | Fev 5<sup>th</sup> | Fev 6<sup>th</sup> | Fev 7<sup>th</sup> | Fev 8<sup>th</sup> | Fev 9<sup>th</sup> | Fev 10<sup>th</sup> | Fev 11<sup>th</sup> |
| $r$ | 0.18725 | 0.19019 | .19480 | 0.19919 | 0.20281 | 0.2072 | 0.21095 |
| $T$ | 102652 | 99103 | 94647 | 91275 | 88993 | 86654 | 85011 |

As it can be seen a reduction in the value of the parameter r causes an increase in the number of the total amount of infected individuals predicted by the model, *T*, and vice-versa. Around February 5*^th^* (exactly, as expected, when the number of new cases start to decrease) we can see an effective decrease in *T*.

## 5. Fitting the model to the data of SÃo Paulo city

São Paulo city is Brazilian financial capital and it was the city where the first case of COVID-19 was confirmed in Brazil. The population of it is around 12 million people which puts it into a very nice position to be compared to Wuhan (11.08 million inhabitants). Besides, it is big enough in several senses to be compared to several countries in Europe. The demographic density of São Paulo though is 7, 398.26 inhabitants/*km*^2^ (according to IBGE’s census [9]) about 5.6 times that of Wuhan.

We have found some very trustable data of the epidemic for the city of São Paulo at the website of the state government (see [6]). Yet we question the values of the number of infected individuals in some specific dates in which they are very discrepant from the ones of the previous and following days. We attribute such distortions to several factors such as under notification, delay on the publication of the screening results as well as weekend and holiday periods. We believe such punctual discrepancies will not cause any problem in our analysis.

We now fit the data for the city of São Paulo, (obtained at [6]) updated to May 8*^th^*. Using again software Mathematica. We get the following function for São Paulo’s total number of infected individuals:

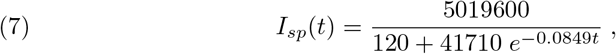

which means *r* = 0.0849 and *T* ≈ 41, 830. We plot the graphic of the above function (considering February 25*^th^* as day 1) together with the official data obtained for São Paulo (see figure 4) to see that the matching is reasonably good:

**Figure 4.**
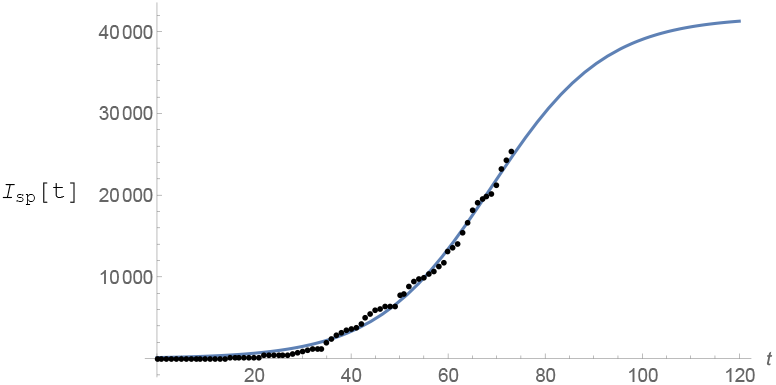
Total number of infected individuals fitting the data for São Paulo (From February 25 *^th^* to May 8*^th^*).

## 6. Avoiding Total Collapse of the Public Health System of SÃo Paulo

### 6.1. Estimating the variation needed in r to relieve the Intensive Care Units of the Public Health System of São Paulo

According to local city authorities by the end of May of 2020, São Paulo shall have 1,440 Intensive Care Unit (ICU) beds in the public hospital network (according to [12]). The mean need for these kind of hospital beds for COVID-19 patients in São Paulo city is about 30% of the number of infected individuals. On May 8*^th^*, São Paulo city had 25, 367 confirmed cases and the average time of intern hospitalization was of about 15 days. With the help of function *I_sp_*(*t*) given by (7) and software Mathematica we have computed when the total number of active infected individuals of a period of 15 consecutive days reaches 3, 840, for then, 30% of it: 1,152 would represent 80% of all the available ICU beds putting under alert the health ICU system. From the website [10] we get that from the total number of infected individuals since the beginning of the epidemic up to day *t*, *I_sp_*(*t*), approximately 7% of it has died and around 40% of it has recovered by day *t*, which leaves around 53% open cases on day *t*. Therefore the total number of active cases on day *t* is *Act*(*t*) = 0.53I_sp_(t). So, the total number of active cases over a period of 15 consecutive days is *A*_0_(*t*) = *Act*(*t*) *− Act*(*t −* 15) =.53(*I_sp_*(*t*) *− I_sp_*(*t −* 15)).

In figure (5) it is shown the period of time such a number of active infected cases is above 3, 840 individuals. According to the model it has been happening since around day 57*^th^* (April 22*^th^*). If non public more strict intervention is taken the total amount of simultaneously infected individual shall grow up to 6, 836 leading the number of total demand for ICU treatment to reach 2, 051 before it starts to decrease, causing total chaos in the system.

**Figure 5.**
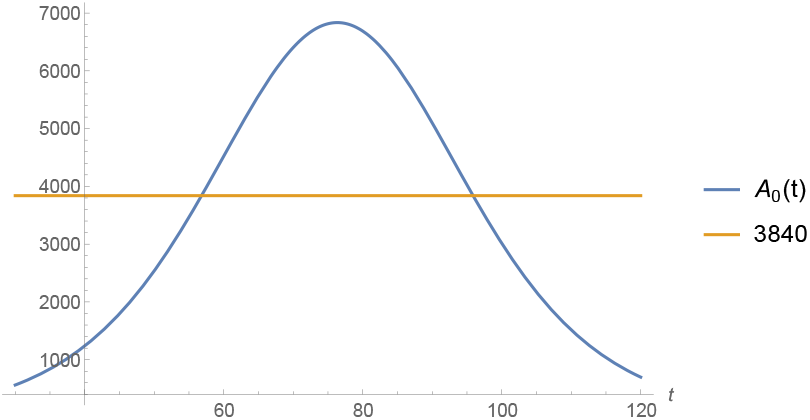
The total number of active infected individuals over a period of 15 days.

We now present a study of how the public policies can help speed up the reduction of such figures. We do our analysis considering the actions as if they were taken on May 8*^th^*, the 73*^th^* day from the beginning of the outbreak of the epidemic in São Paulo. Using function *I_sp_*(*t*) for day 73*^th^* we get the following relation between r and *T*:

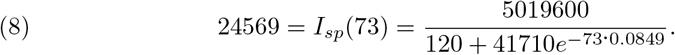

We consider the following function

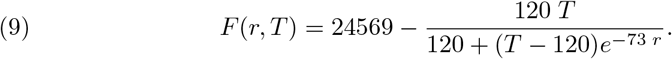

Clearly *F*(*r*_0_ = 0.0849, *T*_0_ = 41830) = 0 and using the Implicit Function Theorem (see Section 8) we can estimate how much a small variation *dr*, in the value of the parameter *r*, can change the value of *T*, the total number of infected individuals during the whole epidemic. If *dr* is small, such corresponding change, *dT*, is well approximated by the following relation:

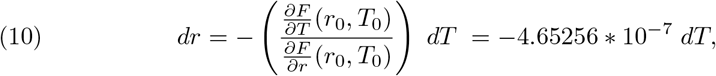

Therefore in order to cause a reduction of, say, 2% in the value of *T* at point *T*_0_ = 41, 830 we need a variation in *r* (at point *r*_0_) of approximately 0.0004, (were this action taken around May 8*^th^*). Because such a variation shall happen at the point *r*_0_ = 0.0849, such a decrease represents a reduction of 0.46% in the value of the parameter r.

### 6.2. Estimating the necessary period of time for an specific variation in the value of *r*

Our last task is to be able to estimate how a variation of 0.46% in the parameter r can be realistic achieved and how long such a process shall take in São Paulo under its current quarantine regime restrictions.

The results of the public policies adopted in Wuhan shall give us a hint. An attentive look at table (1) shows us that the period of time taken for achieving an specific variation on r depends on “ where in the curve we are”. So, what we must do first, is to fit together the Wuhan and the São Paulo curves so that both curves can represent similar events at similar dates and after that quantify how long it took for the Chineses to get the variation we want (0.46% in the value of r) at the corresponding value for *r*_0_. Then, we use it as a baseline for the same reasoning for the case of the city of São Paulo.

In this paper I consider that the first case in Brazil was diagnosed in São Paulo on February 25*^th^* in accordance to the website Worldometer [10] (for the Brazilian government it was on February 26*^th^*). So the epidemic in São Paulo is in approximately 35 days of delay in relation to Wuhan (which first information of infected individuals was of 571 cases and it was done on January *22^nd^* (see [10])). On the other hand the number of infected in São Paulo at time *t*_0_ (day 1) was just 1 individual. Furthermore São Paulo only reached the number of 571 infected individuals (the same number China had on the Chinese day 1) on March 25*^th^*, the 64*^th^* day from the beginning of the outbreak in São Paulo. These differences of initial date and data do cause a big difference in the real dates and in the order of magnitude of the figures concerning Wuhan and São Paulo so that I shall include 2 “adjusting parameters”, *μ*_0_ and *K* in order to better compare both data. The parameter *μ*_0_ will take care simultaneously of the magnitude of the Chinese figures (scaling) as well as the fact that we are considering the data in Wuhan being 80% of that of the whole China. We shall call *Ī_w_* (*t*) = *μ*_0_*I_c_*(*t*), “the adjusted curve of Wuhan” and we shall fit it to the curve of São Paulo, translated *K* days.

I search for a “scaling” of the curve for China in such a way to the adjusted curve to be “a best approximation” for the “translated” curve for São Paulo. The translation we are working with is of a *K* ∊ (35, 60) days based on my previous observations. This approximation should be such that, near day (73 – *K*)*^th^* their values as well as the values of their first derivatives are in “a best approximation”. This demands us to use the norm of the *C*_1_-space, i.e. the space of functions with continuous first derivatives. Such a norm is given by:

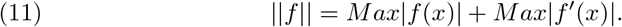

In figure (6) I have plotted the values of the norms of the difference, *I_sp_*(*t* + *K*) − *μ*_0_*I_c_*(*t*) for values of *μ* ∊ (0.33, 0.34) with a step of 0.005 and *K* ∊ (30, 65) with a step of 0.75. After refining such analysis I have found the optimum values to be *μ*_0_ = 0.337 and *K* = 45. In figure (7) I have plotted both graphics to see how well they match for such values of *μ*_0_ and *K* close to day 28*^th^* (73 – 45). We remark that in both figures (6) and (7), in the horizontal axis, the values refer to the corresponding values of *K* ranging from 30 to 65 with intervals of 0.75.

**Figure 6.**
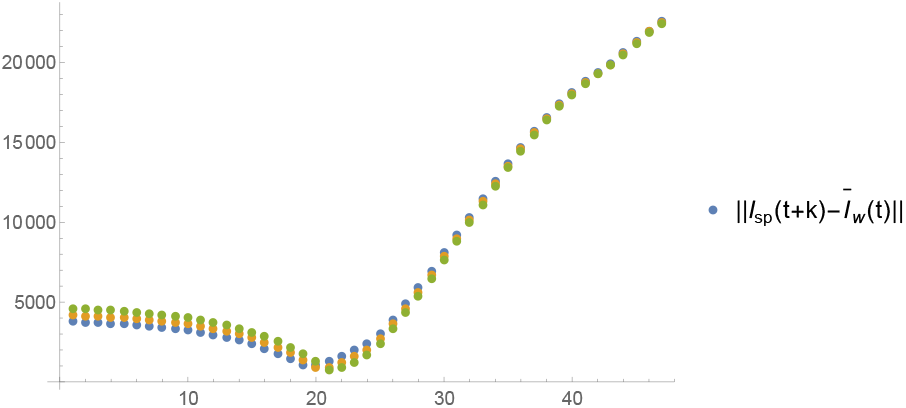
The values of the norms of the difference *I_sp_*(*t* + *K*) − *Ī_w_*(*t*) for several values of *μ* ∊ (0.33, 0.34) and of *K* ∊ (30, 65).

**Figure 7.**
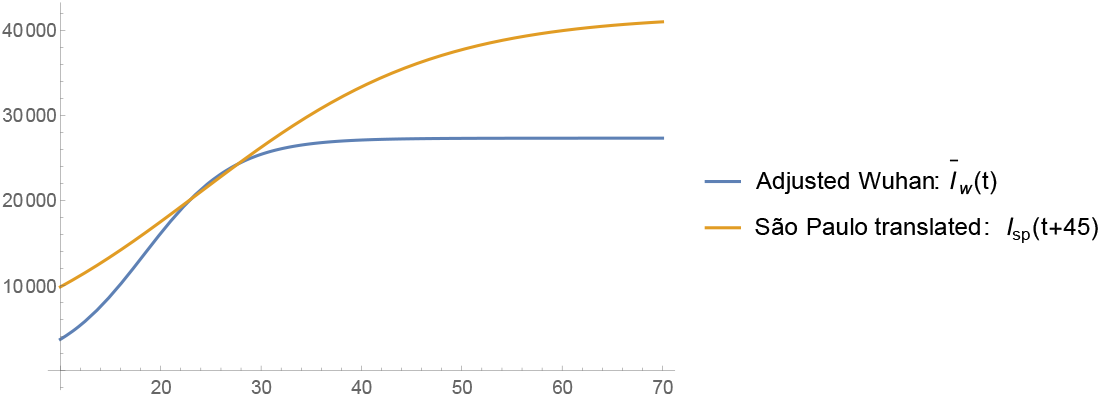
The curves of *I_sp_*(*t* + 45) and *Ī_w_*(*t*).

So, I consider that the graphic of *I_w_*(*t*) = 0.337 *I_c_*(*t*) = 0.42(0.8*I_c_*(*t*)) = 0.42*I_w_*(*t*) represents very well the curve of *I_sp_*(*t*) translated 45 days forwards. I now use it to estimate how long the quarantine regime in Wuhan took to produce a variation of 0.46% in the value of the parameter *r* at day 28*^th^*.

I have computed the values of the parameter *r* relative to the data of Wuhan on days 28*^th^* and 29*^th^* and they were found to be *r*_28_ = 0.202813 and *r*_29_ = 0.20726 respectively which means a variation of approximately 2% on 24 hours, in the value of the parameter r at this position on the curve. I suppose linearity of these relations is the best we could hope for, so under this hypothesis, a variation of 0.46% of the parameter r could not be achieved before 5.5 hours. As I have already pointed out the demographic density of the exposed (non quarantined) susceptible individuals of Wuhan was 8 individuals per *km*^2^ on the other hand the mean level of quarantined portion of the population in São Paulo is accounted to be around 50% of its total population (according to Globo’s television website [7]) which means about 6 million individual wandering over a total area of 1, 622 *km^2^* meaning a demographical density of approximately 3, 700 individuals per *km*^2^; approximately 462.5 times the demographical density of exposed individuals in Wuhan. I use a simple rule of direct proportion to estimate the time spent in the city of São Paulo (under a quarantine of 50% of the total population) to decrease the number of total final infected individuals predicted by the model, i.e., 41, 830, by 2%: 5.5 × 462.5 = 2, 553 hours, i.e. approximately 106 days.

The conclusion is that a variation of 0.46% in the value of *r* in the city of São Paulo, would take 106 days (starting from May 8*^th^*) and that would cause a variation of about 2% in the number of total infected individuals predicted by the model, reducing it from 41, 830 to 40, 994, supposing a quarantine accession of 50% of the whole population of São Paulo city.

I repeat the above analysis for several other cases of quarantine accession and corresponding reductions in *T*_0_ of 2%, 3%, 5% and 7%. I point out that the level of imprecision of our forecasting increases when the percentages of reduction of *T*_0_ increases. In table (2) I present the corresponding amount of days is needed in each one of such studied cases.

Let *A_η_*(*t*) for *η* ∊ {0, 2, 3, 5, 7} denote the functions that represent the total number of active cases over a period of 15 consecutive days (ending on day *t*) after a reduction of *η*% had been obtained in the value of *T*_0_. I conclude this study analyzing the effects the reductions in *T*_0_ can cause on *A_η_*(*t*) for this will have direct impacts on the demand for ICU beds of the public health system of the city of São Paulo. The maximum values of the *A_η_*(*t*) functions decrease with *η* and they happen earlier than for *A*_0_(*t*). Also *A_η_*(*t*) becomes greater than 3840 a little bit earlier, unfortunately (see figure (8)). In table (3) I present the corresponding maximum values of *A_η_*(*t*) for *η* ∊ {0, 2, 3, 5, 7} and the percentage of reduction it represents in relation to the maximum value of *A*_0_(*t*).

**Table 2.** Number of days needed to a reduction of *η*% in the total final number of infected individuals at the end of the epidemic, *T*_0_, under quarantine regime of Δ% of accession.

| $\eta\% \backslash \Delta\%$ | 50% | 55% | 60% | 65% | 70% | 75% | 80% |
| --- | --- | --- | --- | --- | --- | --- | --- |
| 2% | 106 | 95 | 85 | 74 | 64 | 53 | 42 |
| 3% | 159 | 143 | 127 | 111 | 95 | 79 | 63 |
| 5% | 265 | 238 | 211 | 185 | 158 | 132 | 105 |
| 7% | 370 | 333 | 296 | 259 | 222 | 185 | 148 |

**Figure 8.**
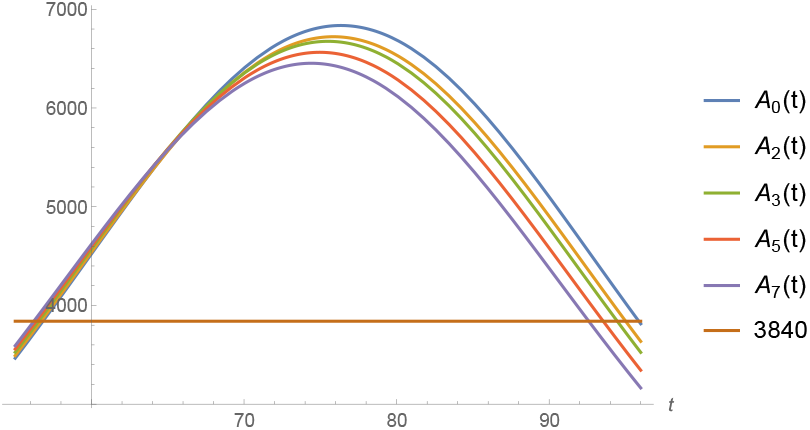
*A_η_*(*t*) for *η* ∊ {0%, 2%, 3%, 5%, 7%} and the level of 3840 representing 80% of the demand for ICU beds.

**Table 3.** The maximum values of *A_η_*(*t*) and the corresponding percentage of reduction it represents from the maximum value of *A*_0_(*t*) under a reduction of *η*% in *T*_0_.

| $\eta \backslash \Delta$ | Max $A_\eta(t)$ | Percentage of Reduction |
| --- | --- | --- |
| 0 % | 6836 | 0 % |
| 2 % | 6722 | 1.66 % |
| 3 % | 6675 | 2.34 % |
| 5 % | 6564 | 3.97 % |
| 7 % | 6453 | 5.6 % |

## 7 Conclusion

We have used the logistic model to represent the COVID-19 epidemic in the city of Wuhan in China with astonishing good fitting. We analyze the effect some non-pharmaceutical measures has had in the epidemic spreading in Wuhan. Specifically we estimate how many days of a 99.5% effective quarantine are necessary to cause a variation in the spreading parameter r of 0.46%.

We use the same model to the ongoing epidemic in São Paulo to forecast its final results. We conclude that if nothing else be done the number of infected people in São Paulo will reach around 40,000 before the end of May.

Based on the official number of ICU beds in the public health system we mathematically estimate the amount of variation in the parameter *r* which is necessary to cause a decrease in the value of the total number of simultaneously infected individual in need of special intensive care in order to guarantee this number remains below 80% of the total capacity of ICU beds of the public health system. Then, based on the experience in Wuhan, we estimated how long it would take for the city of São Paulo under several different level of accession to the quarantine, to produce this desired variation in the parameter r.

## Data Availability

All the data referred to in the manuscript is either included in the manuscript of referred to in the bibliography section. All the website accessed are also referred in the bibliography section.

## 8. Appendix

We present the Implicit Function Theorem used in Section 6:

### Theorem 1

*Suppose F*: ℝ^2^ → ℝ *is a continuously differentiable function defining a curve F*(*x*, *y*) = 0. *Let* (*x*_0_, *y*_0_) *be a point on the curve such that F*(*x*_0_, *y*_0_) = 0 *and* 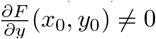 *then for the curve around* (*x*_0_, *y*_0_) *we can write y* = *f*(*x*), *where f is a real differentiable function. Furthermore the derivative of such function is given by:*

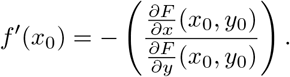

## CONFLICT OF INTERESTS

The author declares he has no conflict of interests.

## Notes

### Competing Interest Statement

The authors have declared no competing interest.

### Funding Statement

no external funding was received

### Author Declarations

not necessary in the present case

